# Effects of the COVID-19 pandemic on individuals with Chemical Intolerance

**DOI:** 10.1101/2024.05.23.24307835

**Authors:** Raymond F. Palmer, David Kattari, Monica Verduzco-Gutierrez

## Abstract

**Background:** The Center for Disease Control has estimated that over 24 million have been infected with COVID-19 in the US with over 6,700,000 being hospitalized, and over 1,174,000 deaths. Several other industrialized countries show similar numbers (CSSE, 2021). Chemical Intolerance (CI) is characterized by multi-system symptoms initiated by a one-time high dose or persistent low-dose exposure to environmental toxins including chemicals, foods and drugs. With an estimated 20% prevalence in the US, the symptoms of CI include fatigue, headache, weakness, rash, mood changes, musculoskeletal pain, gastrointestinal issues, difficulties with memory, concentration, and respiratory problems which are similar to COVID-19 and its sequelae. The purpose of this study was to determine if the pandemic had differential effects on those individuals with CI.

**Methods:** A large U.S. population-based survey was launched involving 7,500 respondents asking if they ever had COVID-19, what the severity of it was, and if they have long COVID-19. Respondents were also assessed for CI using the Quick Environmental Exposure and Sensitivity Inventory (QEESI), a 50-item validated questionnaire designed to assess intolerances to inhaled chemicals, foods, and/or drugs. Respondents were classified as Low, Medium, or High CI.

**Results:** Those in the High Chemical Intolerance class reported a greater COVID-19 prevalence, symptom severity, and long COVID-19 then in the Medium and Low CI classes (P<.0001). These associations were independent of race, ethnicity, income, age, and gender. However, there was significantly increased odds of COVID-19 severity among females and those over 45 years old. Asian individuals were least likely to have severe symptoms compared to White individuals (OR = 0.60). Black/African American individuals reported a lower prevalence of COVID-19 than Non-Hispanic Whites (NHW), but African American individuals with high CI reported 2.2 greater odds of reporting COVID-19 prevalence. Further, African American individuals had significantly greater odds of increased symptom severity.

**Discussion:** Prior studies showed that higher risk for COVID-19-19 infection include the elderly, male sex, those with pre-existing comorbidities (e.g., challenged immunities) and those from minoritized racial/ethnic groups. The results of this study suggest that those with CI be included in a high risk group. Various risk subsets may exisit and future investigations could identify different risk subsets. Understanding these subgroups would be helpful in mounting targeted prevention efforts.

## 1.0 Introduction

The SARS-CoV-2 virus that causes COVID-19 infection first emerged in December 2019. By March 2020, the World Health Organization declared the outbreak a global pandemic [1]. The CDC has estimated that 78% of Americans have been infected at least once and that 97% of adults had antibodies to the virus from either infection, vaccination, or a combination of the two [2]. Estimates to date indicate that over 24 million have been infected in the US with over 6,700,000 being hospitalized, and over 1,174,000 deaths [2,3]. Several other industrialized countries show similar numbers [3].

Those infected with COVID-19 have a considerable heterogeneity of symptoms, from no symptoms at all to mild to severe symptoms. Symptoms usually last several days after exposure and can include fever, cough, shortness of breath or difficulty breathing, chills, fatigue, muscle pain, body aches, headache, sore throat, new loss of taste or smell, congestion, runny nose, nausea, vomiting, and/or diarrhea [4].

Persistent symptoms lasting three or more months after the initial infection is referred to as Long COVID-19, with persistent long-term health issues involving cardiovascular, pulmonary, renal, neurologic, and psychological functions. [5,6]. “Brain fog” (unusual forgetfulness, word finding deficits, and inability to concentrate) has been cited as a “particularly frustrating persistent symptom” [6]. It is estimated that up to 13% of those infected with COVID-19 will develop long COVID-19, and account for 30% of COVID-19 related hospitalizations [7].

Individuals with the greatest risk of serious illness include the elderly, those with chronic medical conditions such as diabetes, cancer, heart disease, and those with weakened immune systems [8,9]. Individuals with Chemical Intolerance (CI) may also be especially vulnerable to infection and to have more severe symptoms. As such, not only do people with CI need to take extra measures to avoid exposure to COVID-19, but they have remained vulnerable when workplaces and public spaces were re-opened, and disinfectant chemicals were widely used to sanitize the environment [6]. The purpose of this paper is to investigate the association between CI and COVID-19.

### 1.1 Chemical Intolerance

CI, also known as Multiple Chemical Sensitivity (MCS), is characterized by multi-system symptoms initiated by a one-time high dose or persistent low-dose exposure to environmental toxins [10]. New-onset intolerances may occur when an individual is subsequently exposed to structurally unrelated chemicals, foods, and/or drugs [10,11]. Similar to the symptoms of COVID-19, the symptoms of CI include fatigue, headache, weakness, rash, mood changes, musculoskeletal pain, gastrointestinal issues, difficulties with memory, concentration, and respiratory problems [12–15].

Assessing CI most often involves the use of the Quick Environmental Exposure and Sensitivity Inventory (QEESI), a 50-item validated questionnaire designed to assess intolerances to inhaled chemicals, foods, and/or drugs. The QEESI offers high sensitivity and specificity that differentiates CI individuals from the general population [16–18]. The QEESI has been well received by the international scientific community [15] with several language translations used in over 16 countries around the world in over 90 publications (see www.TILTreseaarch.org for a comprehensive list).

Estimates of CI prevalence in separate population-based surveys differ by whether it is clinically diagnosed (0.5–6.5%), or self-reported (averaging ∼ 20%) [19–23]. Two nationally representative U.S. population surveys, conducted in 2002 and 2006 [24,25], found a prevalence of 11.1% and 11.6% (respectively) for self-reported chemical sensitivity and 2.5% and 3.9% medically diagnosed MCS. More recent nationally representative U.S. population studies conducted in 2016 found a prevalence of 25.9% self-reported chemical sensitivity and 12.8% reported medically diagnosed “multiple chemical sensitivities” or MCS (Steinemann, 2018). Based on these data, the prevalence of chemical sensitivity may have increased by over 200%, and diagnosed MCS by over 300%, in the past decade [26]. Further, Hojo et al., 2018 provide evidence of similar prevalence increases over a 10-year period in Japan. Using the QEESI they report that the scores for chemical intolerances, other intolerances, and life impacts increased significantly. These increases may be attributed to modern lifestyle exposures and industrialized food consumption [18].

Increasing numbers of patients and researchers attribute CI to well-defined exposure events, such as indoor air contaminants (e.g., fragranced personal care and cleaning products), exposures to pesticides, new construction or remodeling, or a flood- or water-damaged building resulting in mold and bacterial growth (12, 25, 27, 28).

### 1.2 Study purpose

At the beginning of the COVID-19 pandemic (6/2020), a population survey called the Personal Exposure Inventory (PEI 1) was launched. [22]. The prevalence of CI was reported to be 19.3% - commensurate with other population estimates in the literature. Two years later in June 2022, the PEI2 current survey was launched involving questions concerning the impact of COVID-19. (see Figure 1, CDC 2024). In this manuscript, we report the results of the PEI2 investigating the following research questions and corresponding hypotheses to determine if the pandemic had differential effect on those individuals with CI.

1. Did the overall prevalence rate of reported CI increase from PEI1 to PEI2?

**Figure 1.**
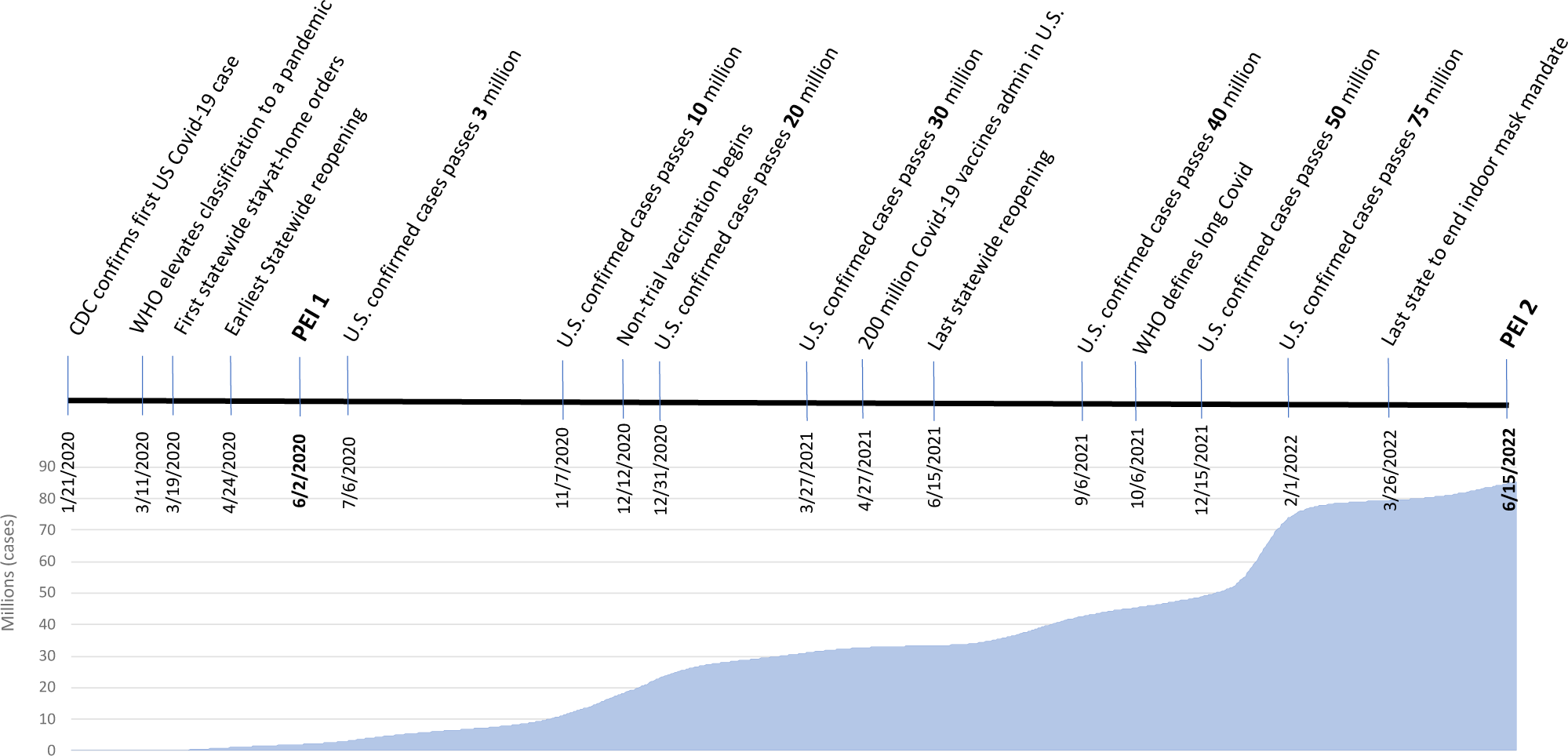
Timeline and trajectory of COVID-19 and our population-based surveys.

*Hypothesis 1*: The prevalence of CI will be higher in PEI2 compared to PEI1.

1) 2. Did those with CI report higher COVID-19 rates than those without CI?

*Hypothesis 2*: The prevalence of CI will be higher among those with CI.

1) 3. Are those with CI more severely affected by COVID-19?

*Hypothesis 3*: Those with CI will be more severely affected by COVID-19.

1) 4. Are those with CI more likely to experience Long COVID-19?

*Hypothesis 4*: The prevalence of CI will be higher among those with CI.

1) 5. Are those with CI more likely to experience reactions to the COVID-19 vaccine?

*Hypothesis 5*: The prevalence of vaccine reactions will be higher among those with CI.

In addition, the effects age, ethnicity, race and gender associated on COVID-19 outcomes are analyzed.

## 2.0 Methods

This observational study involved a population-based survey of U.S. adults aged 18 years and older.

SurveyMonkey recruitment procedures are available here: www.surveymonkey.com. The survey was deployed on 6/15/2022 and ended 6/25/2022. There were 7504 respondents that were randomly selected from nearly 3 million online users of the SurveyMonkey platform. The survey had an abandonment rate of 12.6% and took an average of 12.5 minutes to complete.

The modeled error estimate for this survey was ±1.189%. Respondents were selected from online panels based on the population sizes of all 50 states plus the District of Columbia, as well as by sex, age, race/ethnicity, and educational level within each census region to match the U.S. Census Bureau’s 2015 American Community Survey (ACS) targets.

We had previously deployed a similar survey on 6/1/2020. Details of that earlier survey are previously described [22]. In the current study, respondents were classified into high, medium, or low CI groups, and self-reported COVID-19 prevalence, severity, and long COVID-19. Note that the associations identified in this study are not considered causal.

### 2.1. Survey

Respondents answered an 80-item survey called the Personal Exposure Inventory 2 (PEI 2), which included items concerning individuals’ demographics, medical diagnoses, and CI. Age and income were captured as part of SurveyMonkey’s panel. Age was reported as a four-level categorical variable, with age increasing roughly every 15 years. Income was reported as a ten-level categorical variable, with income increasing by roughly USD 25,000 per level. Race and ethnicity were collected following federal guidelines.

Respondents were asked three primary questions to assess COVID-19 infection: “Have you been diagnosed with, or do you think you have had COVID-19?” Respondents could answer one of five responses:

1) “Yes, I believe I have had COVID-19 (no test),” 2) “Yes, I tested positive for COVID-19 with a home test,” 3) “Yes, I tested positive for COVID-19 by a health professional,” 4) “No,” and 5) “I am not sure.” The second question was: “If yes, please rate the severity of your COVID-19 symptoms on a 0-10 scale. [0 = no symptoms] [5 = moderate symptoms] [10 = disabling symptom].” The third question was “Did your COVID-19 symptoms last for more than 12 weeks (long COVID-19)?” We also ask about vaccination status and if there were reactions to the vaccine (See Appendix 1).

### 2.2. Chemical intolerance assessment using QEESI classification scoring

CI was assessed using the QEESI Chemical Exposures and Symptoms scales [16,17]. These scales are used to classify participants into CI severity groups [16,17]. Each scale contains 10 items that are rated from 0 to 10 on a Likert scale: 0 = “not at all a problem” to 10 = “severe/disabling symptoms.”. Total scores for each scale range from 0 to 100. The cut-off criteria for “High CI” are scores greater than or equal to 40 on both the Chemical Exposures and Symptoms scales. This is regarded as “very suggestive” of CI. Scores from 20 to 39 on one or both scales are considered “Medium CI.” Scores less than 20 on both scales are classified as “Low CI”.

### 2.3. Data Quality Control Checks

The 7504 survey records were assessed for data quality (DQ) encompassing completeness, validity, or accuracy concerns; six measures were required to exclude all surveys indicating one or more DQ concerns. Records with these concerns were excluded from the analytic data set. Figure 2 depicts the flow of data exclusions leading to the final analytic dataset. Some of the DQ measures might technically be accurate (e.g., “male and breast implants”), but with an abundance of caution, they were excluded. The same could be said for the survey time measure: with a survey containing a minimum of 53 questions, it is unlikely that a respondent could read and respond accurately to all questions in under three minutes. By omitting any records that violated one or more DQ measures, 2170 records were excluded (28.9%). We have taken this approach to help ameliorate some well-known DQ concerns associated with web-based surveys, including response probabilities and biases [29,30]. After applying both the data quality, our final analytic sample was N = 5334 (see Figure 2).

**Figure 2.**
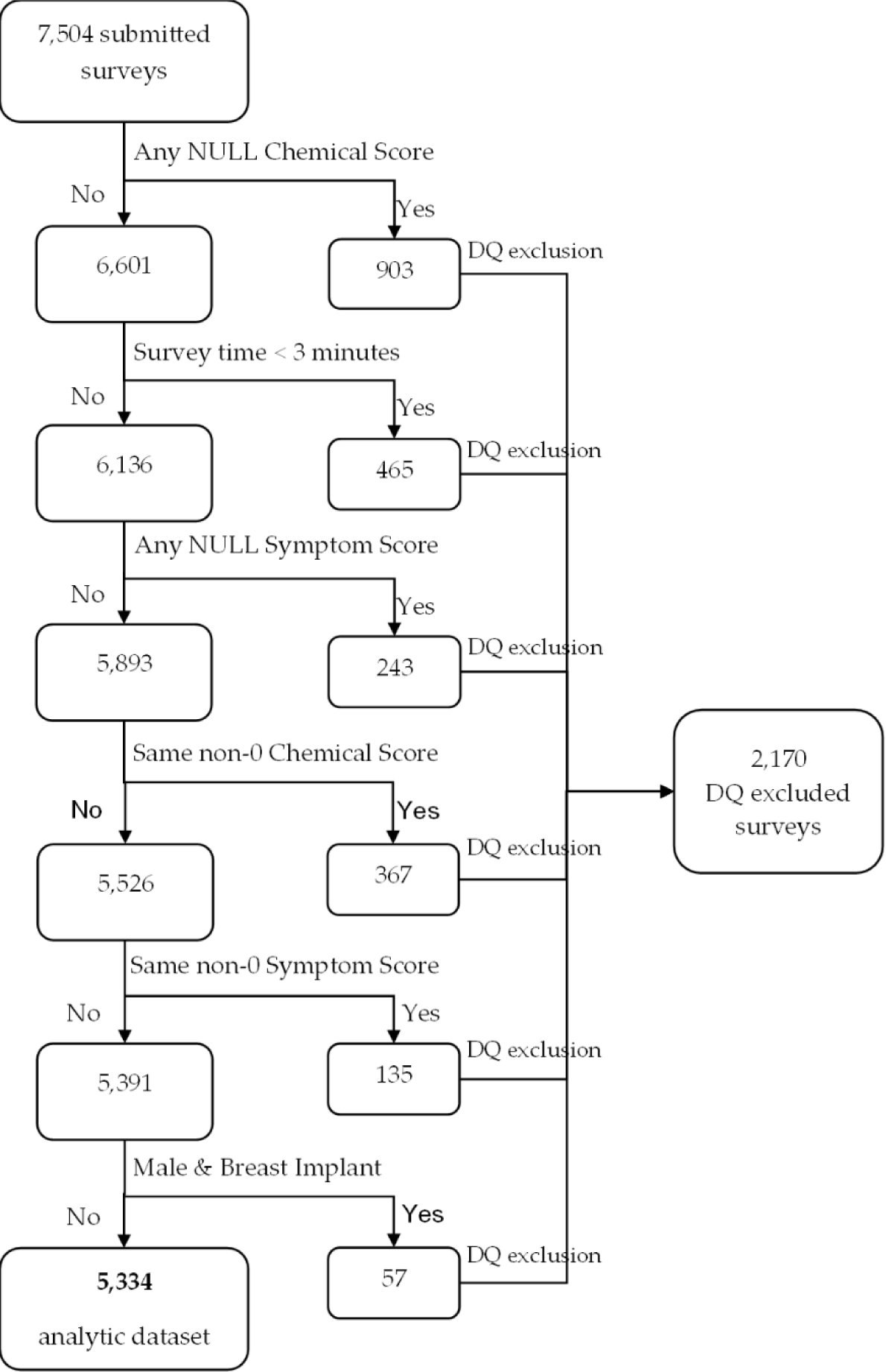
Total survey sample size and final analytic sample after Data Quality Assessment.

### 2.4. Statistical Modeling

COVID-19 Prevalence: A binary logistic regression was conducted to determine if there was a relationship between CI and COVID-19 prevalence. The binary dependent variable, “Reported COVID-19,” was defined as any positive response to the question pertaining to having been diagnosed with COVID-19. We included six independent variables: CI as defined by QEESI (low, medium, high), Sex, Age, Income, Race, and Ethnicity.

#### Long COVID-19

A similar logistic regression was conducted to determine if there was a relationship between CI and Long COVID-19. The binary dependent variable, “Long COVID-19,” was defined as a positive response to the question “Did your COVID-19 symptoms last for more than 12 weeks (long COVID-19)?” We also include the aforementioned CI and covariates.

#### COVID-19 Severity Score

A Standard Least Squares linear regression was conducted to determine the relationship between CI and COVID-19 severity. The dependent variable was normally distributed and ranged between 0 and 10. We included six independent variables: CI as defined by QEESI (low, medium, high), Sex, Age, Income, Race, and Ethnicity.

#### Odds Ratios for COVID-19 severity

Was furthered modeled as a logistic regression with the binary dependent variable as any COVID-19 severity score equal to or greater than 8. We also conducted a sensitivity analysis on cut points between COVID-19 severity scores of 5+ and 8+. Odds ratios varied little between the candidates with the 8+ cut point demonstrating significant effects for Age and Race. The 8+ cut point further fit the data and reduced the critical population to 22.6% (only the most severe reports of COVID-19).

A p-value of 0.05 and 95% confidence intervals were used to determine statistical significance for all models. Analyses were conducted using SAS and JMP statistical software [31,32]. Respondents needed to be at least 18 years old. Participation consent was obtained online before survey was administered. This research program was approved by the University of Texas Health Science Center at San Antonio Internal Research Board protocol number 20220246EX.

## 3.0 Results

Table 1 depicts the distribution of the study variables. Non-Hispanic White respondents were more prevalent in the sample than other ethnic or racial group. Nearly one third (31.3%) of the sample reported having tested positive for COVID-19. The percentage increases to 42.7 by including those who report they believed they had COVID-19 but were not tested.

**Table 1.**
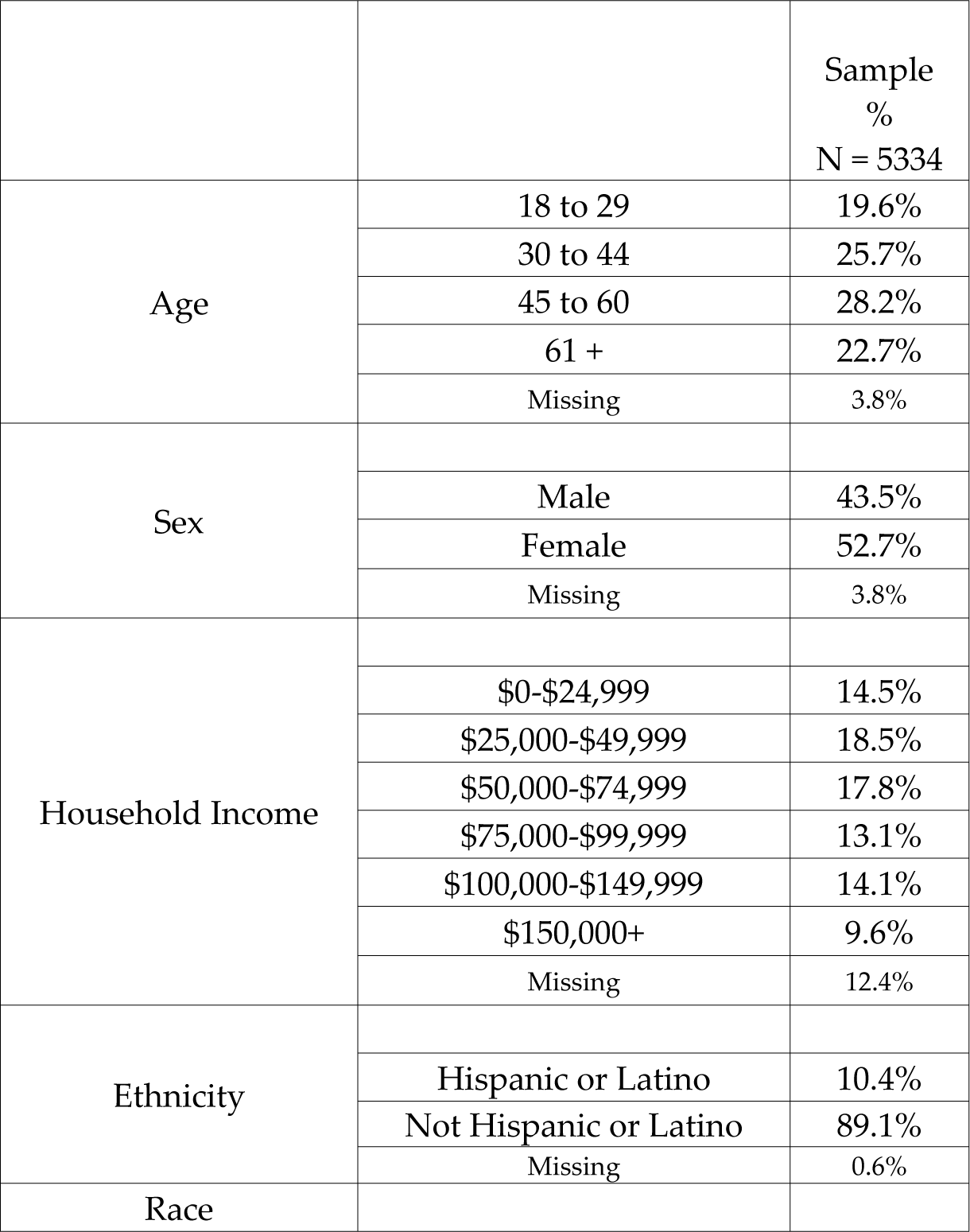
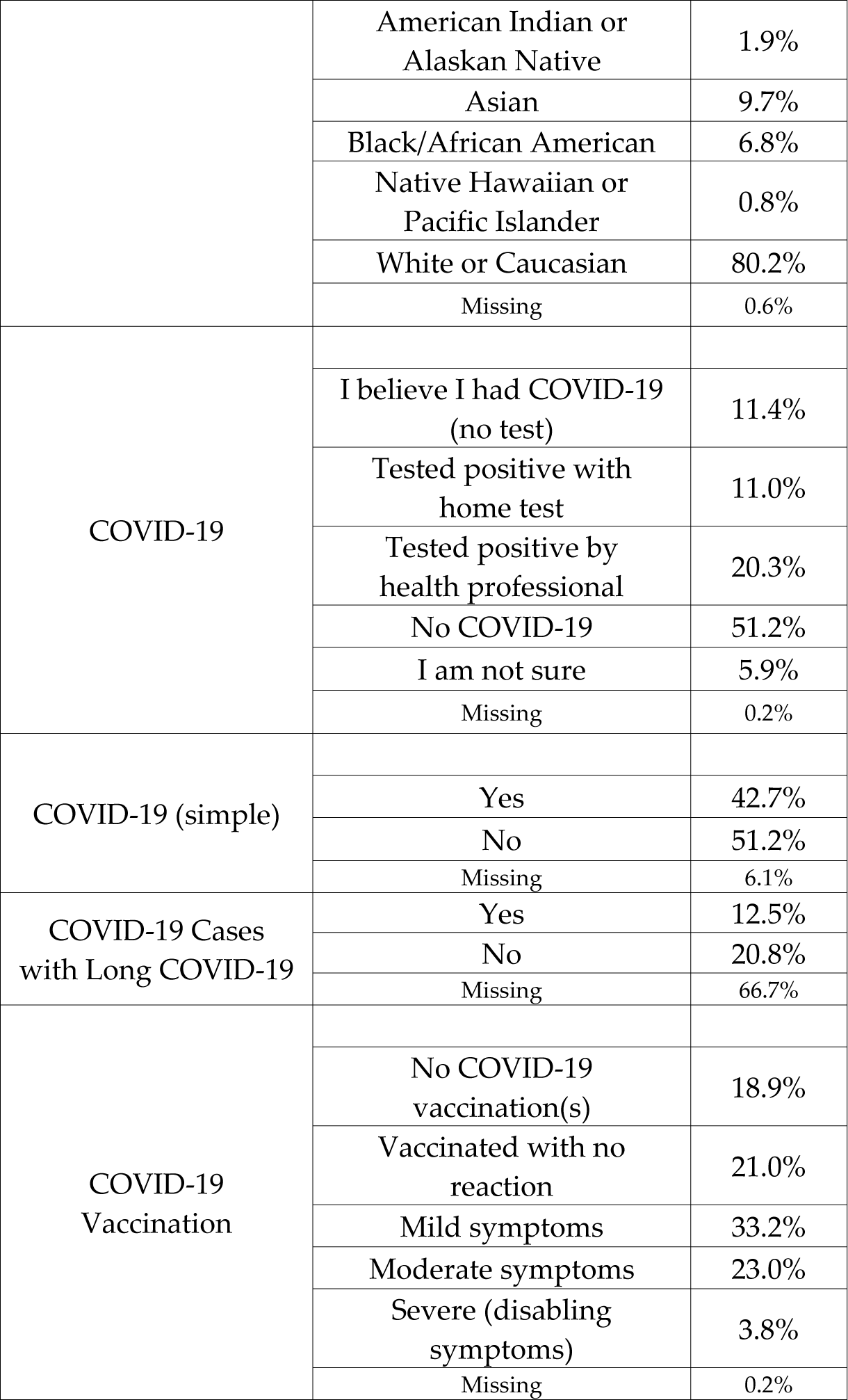
Sample Demographics.

|  |  | Sample<br>%<br>N = 5334 |
| --- | --- | --- |
| Age | 18 to 29 | 19.6% |
|  | 30 to 44 | 25.7% |
|  | 45 to 60 | 28.2% |
|  | 61 + | 22.7% |
|  | Missing | 3.8% |
| Sex | Male | 43.5% |
|  | Female | 52.7% |
|  | Missing | 3.8% |
| Household Income | \$0-\$24,999 | 14.5% |
| | \$25,000-\$49,999 | 18.5% |
| | \$50,000-\$74,999 | 17.8% |
| | \$75,000-\$99,999 | 13.1% |
| | \$100,000-\$149,999 | 14.1% |
| | \$150,000+ | 9.6% |
|  | Missing | 12.4% |
| Ethnicity | Hispanic or Latino | 10.4% |
|  | Not Hispanic or Latino | 89.1% |
|  | Missing | 0.6% |
| Race |  |  |
|  | American Indian or Alaskan Native | 1.9% |
|  | Asian | 9.7% |
|  | Black/African American | 6.8% |
|  | Native Hawaiian or Pacific Islander | 0.8% |
|  | White or Caucasian | 80.2% |
|  | Missing | 0.6% |
| COVID-19 |  |  |
|  | I believe I had COVID-19 (no test) | 11.4% |
|  | Tested positive with home test | 11.0% |
|  | Tested positive by health professional | 20.3% |
|  | No COVID-19 | 51.2% |
|  | I am not sure | 5.9% |
|  | Missing | 0.2% |
| COVID-19 (simple) |  |  |
|  | Yes | 42.7% |
|  | No | 51.2% |
|  | Missing | 6.1% |
| COVID-19 Cases with Long COVID-19 | Yes | 12.5% |
|  | No | 20.8% |
|  | Missing | 66.7% |
| COVID-19 Vaccination |  |  |
|  | No COVID-19 vaccination(s) | 18.9% |
|  | Vaccinated with no reaction | 21.0% |
|  | Mild symptoms | 33.2% |
|  | Moderate symptoms | 23.0% |
|  | Severe (disabling symptoms) | 3.8% |
|  | Missing | 0.2% |

Table 2. shows the study variables by CI class. All percents are presented as row percents. Compared to the Low CI group, those in the High CI group include significantly fewer individuals over 60, incomes under 25k per year, and Female sex. Further, 43.5% of Hispanic respondents where in the High CI class compared to 29% of Non-Hispanic White respondents. Black and Asian respondents reported significantly higher rates of CI compared to NHW.

**Table 2.** Study variables by Chemical Intolerance category.

| Study Variables* | Response Categories | Low CI | Mid CI | High CI |
| --- | --- | --- | --- | --- |
|  |  | N= 864<br>(16.2 %) | N= 2863<br>(53.7 %) | N= 1607<br>(30.1 %) |
|  | 18 to 29 | 14.2% | 53.9% | 31.8% |
|  | 30 to 44 | 13.2% | 50.1% | 36.7% |
|  | 45 to 60 | 12.2% | 53.9% | 33.9% |
|  | 61 + | 24.4% | 56.0% | 19.6% |
|  | *Missing* | 27.1% | 61.1% | 11.8% |
| Sex |  |  |  |  |
|  | Male | 19.7% | 51.5% | 28.8% |
|  | Female | 12.5% | 55.0% | 32.6% |
|  | *Missing* | 27.1% | 61.1% | 11.8% |
| Household Income |  |  |  |  |
| | \$0-\$24,999 | 9.2% | 55.0% | 36.4% |
| | \$25,000-\$49,999 | 14.0% | 55.0% | 31.0% |
| | \$50,000-\$74,999 | 17.7% | 51.3% | 30.9% |
| | \$75,000-\$99,999 | 16.1% | 54.0% | 29.9% |
| | \$100,000-\$149,999 | 16.8% | 53.7% | 29.4% |
| | \$150,000+ | 19.3% | 46.7% | 34.1% |
|  | *Missing* | 22.4% | 59.2% | 18.4% |
| Ethnicity |  |  |  |  |
|  | Hispanic or Latino | 11.6% | 45.0% | 43.5% |
|  | Not Hispanic or Latino | 16.7% | 54.7% | 28.6% |
|  | *Missing* | 20.0% | 60.0% | 20.0% |
| Race |  |  |  |  |
|  | American Indian/Alaskan Native | 10.9% | 46.5% | 42.6% |
|  | Asian | 12.6% | 51.8% | 35.6% |
|  | Black/African American | 12.1% | 46.4% | 41.5% |
|  | Native Hawaiian/ Pacific Islander | 19.1% | 52.4% | 28.6% |
|  | White or Caucasian | 17.1% | 54.7% | 28.3% |
|  | *Missing* | 20.0% | 60.0% | 20.0% |
| COVID-19 |  |  |  |  |
|  | I believe I had COVID-19 (no test) | 10.2% | 53.6% | 36.1% |
|  | Tested positive with home test | 11.9% | 51.9% | 36.2% |
|  | Tested positive by health professional | 15.2% | 51.7% | 33.2% |
|  | No COVID-19 | 19.2% | 55.3% | 25.5% |
|  | I am not sure | 13.3% | 50.0% | 36.7% |
|  | *Missing* | 20.0% | 40.0% | 40.0% |
| COVID-19 (simple) |  |  |  |  |
|  | Yes | 13.0% | 52.2% | 34.8% |
|  | No | 19.2% | 55.3% | 25.5% |
|  | *Missing* | 13.5% | 49.7% | 36.8% |
| COVID-19 Symptom Severity | Rated 0=no symptoms, 10=disabling<br>Mean and standard deviation | 4.0 (SD) | 5.1 (SD) | 6.3 (SD) |
| COVID-19 Cases with Long COVID- | Yes | 8.2% | 14.5% | 34.7% |
|  | No | 78.8 | 73.4 | 52.1 |
| 19 | *missing* | 12.9 | 12.1 | 13.2 |
| COVID-19<br>Vaccination |  |  |  |  |
|  | No COVID-19 vaccination(s) | 13.8% | 55.4% | 30.8% |
|  | Vaccinated with no reaction | 19.7% | 55.4% | 25.0% |
|  | Mild symptoms | 18.7% | 55.1% | 26.2% |
|  | Moderate symptoms | 10.6% | 49.2% | 40.2% |
|  | Severe (disabling symptoms) | 9.2% | 42.7% | 48.2% |
|  | *Missing* | 25.0% | 43.8% | 31.3% |

\*Comparisons between groups across all study variables were significantly different at $P < .0001$ .

### 3.1 COVID-19 Prevalence, Hypothesis 1

This hypothesis tested if the survey estimates of CI administered in 2022 were higher than our baseline survey CI estimates taken in 2020. The prevalence of QEESI criteria for CI significantly increased from 20.6% to 30.1% between the 2-year study period (p < .0001) (see fig 1).

### 3.2 COVID-19 Prevalence by CI group, Hypothesis 2

We found that 49% of those in the highest CI group reported ever having COVID-19, whereas those reporting low CI group 34% (P <. 0001). This represents an odds ratio of 1.8 for the high CI group compared to the Mid group (95% confidence interval = 1.5 – 2.2); and an OR of (95% confidence interval = 2.2 – 4.9) comparing the High to Low CI group (see Figure 3).

**Figure 3.**
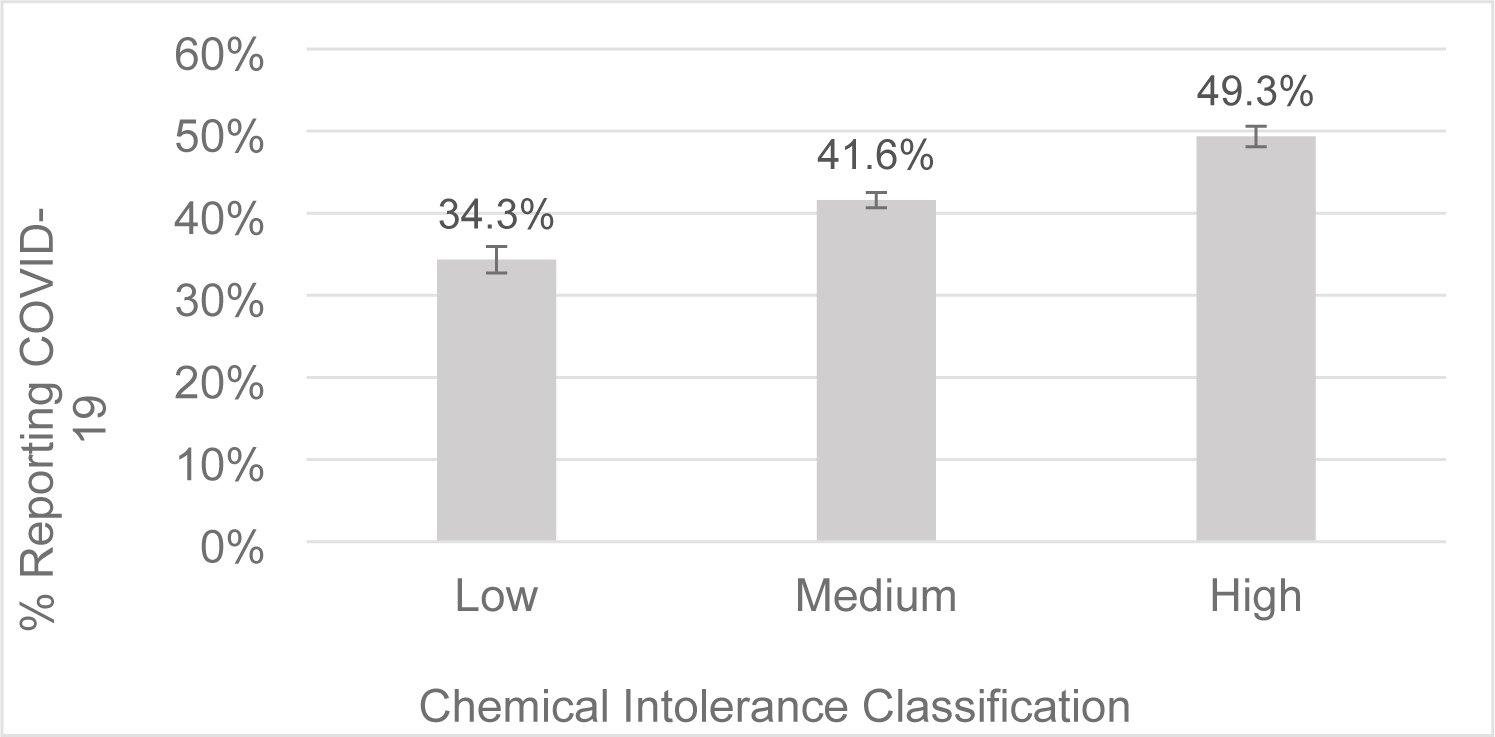
Percent Reporting COVID-19 by Chemical Intolerance Classification.

Forty Eight percent (48%) of those younger than 60 years old reported having COVID-19. In contrast, only 26% who were older than 60 reported COVID-19— corresponding to an OR of 2.4 (95% confidence interval = 2.1 – 2.8).

Hispanic/Latino respondents reported 1.6 higher odds of COVID-19 than non-Hispanic or Latino respondents. Younger respondents reported 2.5 higher odds of COVID-19 than older respondents. High CI respondents reported 3.3 higher odds of having COVID-19 than Low CI respondents. Black or African American respondents reported 0.66 lower odds (33% lower) of COVID-19 than non-Black/African American respondents.

Testing for interactions between CI groups and race, we found a significant interaction among the Black/African American group. Black/African American respondents with High CI show a 2.2 higher odds of reporting COVID-19 than non-Black or African Americans with High CI. (95% confidence interval 1.6 – 3.2).

The final logistic model described above demonstrated good fit to the data (lack of fit index x^2^ p =.10).

### 3.3 COVID-19 Severity, Hypothesis 3

The COVID-19 Severity outcome measure ranged from 0 (none), 5 (Moderate), and 10 (Severe disabling symptoms). With a mean score of 5.3 (standard deviation = 2.6) and an acceptable normal distribution.

OLS regression was run on the sample who reported having COVID-19 (n = 2044). CI categories were the strongest predictor of COVID-19 severity. Each increment from Low, to Mid, to High CI corresponded to a increase in severity score (p < .0001). This can be observed in Figure 4.

**Figure 4.**
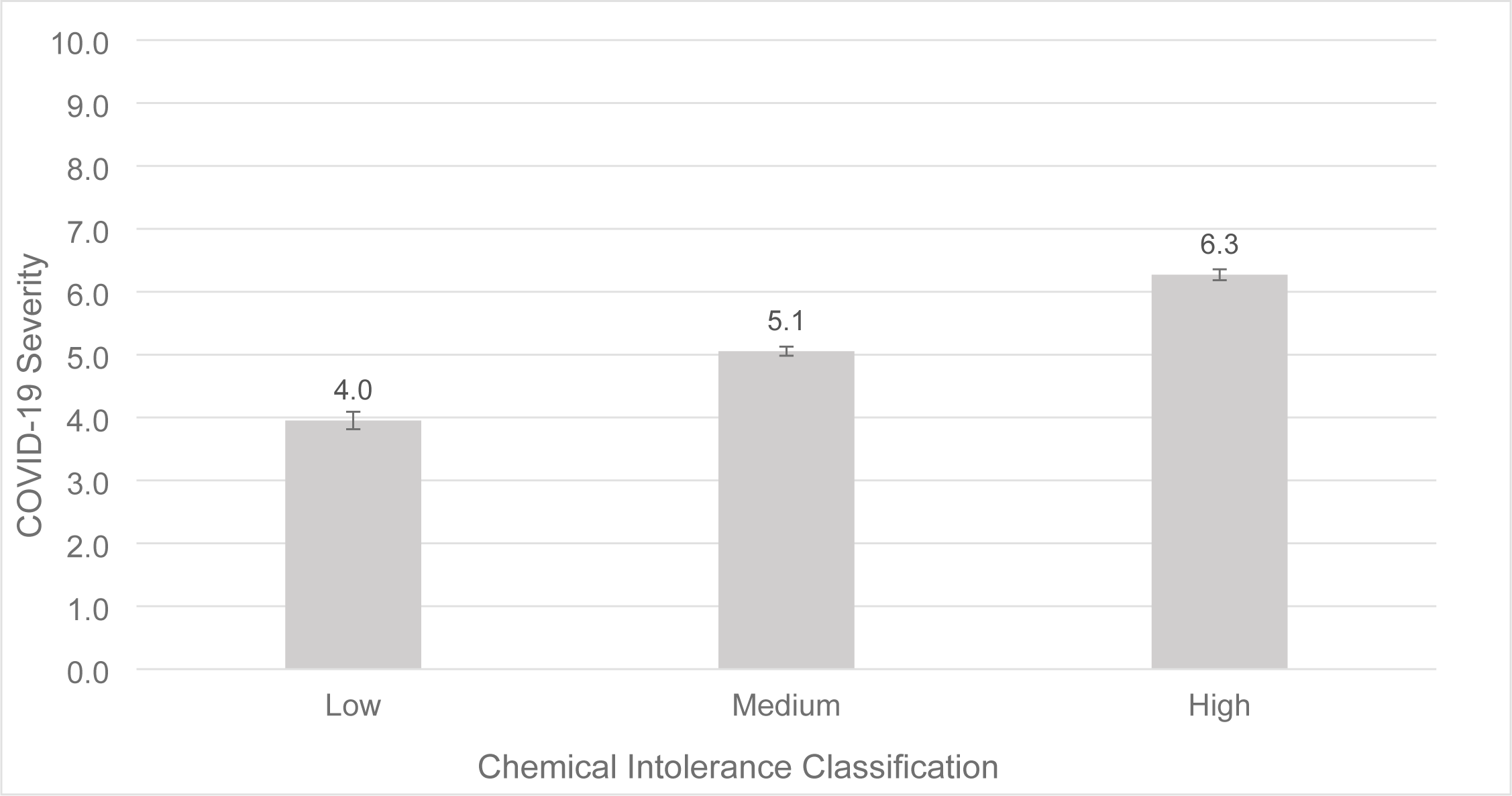
COVID-19 severity by chemical intolerance classification.

Males had a 0.23 lower COVID-19 Severity score than females (p < .001). Those 60 or older had a 0.16 higher COVID-19 Severity scores than younger respondents (p < .05). Model R^2^ was 0.10 with a good lack of fit index (p = 0.37). Ethnicity, Income, and Race were not significant predictors of the COVID-19 severity score.

We also tested various logistic regression models for severity, where severity was dichotomized into either 5 and above (scored 1) and below 5 (scored = 0). A sensitivity analysis was done with other severity cutoff points above moderate severity (6, 7, and 8 cutoffs). Table 3 summarizes these results. In all models, those with High CI had at least 5 times the likelihood of COVID-19 severity above the 8+ cut point compared to those with low CI. Those with Mid CI had at least 2.2 times the likelihood of COVID-19 severity above the 8+ cut point compared to those with low CI.

**Table 3.** Logistic regression models of different COVID-19 severity cut points.

|  | COVID-19 Severity Score Cut Points for logistic model |  |  |  |
| --- | --- | --- | --- | --- |
|  | 5+ | 6+ | 7+ | 8+ <sup>a</sup> |
| High vs. Low CI | 4.5*** | 6.0*** | 5.3*** | 5.0*** |
| Medium vs. Low CI | 1.7*** | 1.9*** | 2.0*** | 2.5*** |
| Female Gender | 1.5*** | 1.3*** | Ns | 1.3*** |
| Age 45 | ns | ns | Ns | 1.3*** |
| Race | ns | ns | Ns | 0.6*** |
| Model Lack of fit (LoF) | 0.002 | 0.005 | 0.004 | 0.23+ |
| N | 1473<br>(64.7%) | 1083<br>(47.6%) | 780<br>(34.3%) | 516<br>(22.6%) |

<sup>a</sup> The model with 8+ severity cutoff is the best fitting model to the data.
\*\*\*P < .0001; ns = not significant

### 3.4 Long COVID-19, Hypothesis 4

QEESI Class, Ethnicity and Race were significant predictors of Long COVID-19. Gender, Income, and Age were not significant predictors.

Respondents in the high CI class had 7.7 times the odds of reporting Long COVID-19 relative to Low classification (95% confidence interval 5.5 – 10.9). Respondents with Mid CI classifications had 2.8 times the odds of reporting Long COVID-19 relative to Low CI classification (95% confidence interval = 2.3 – 3.3) (See Figure 5).

**Figure 5.**
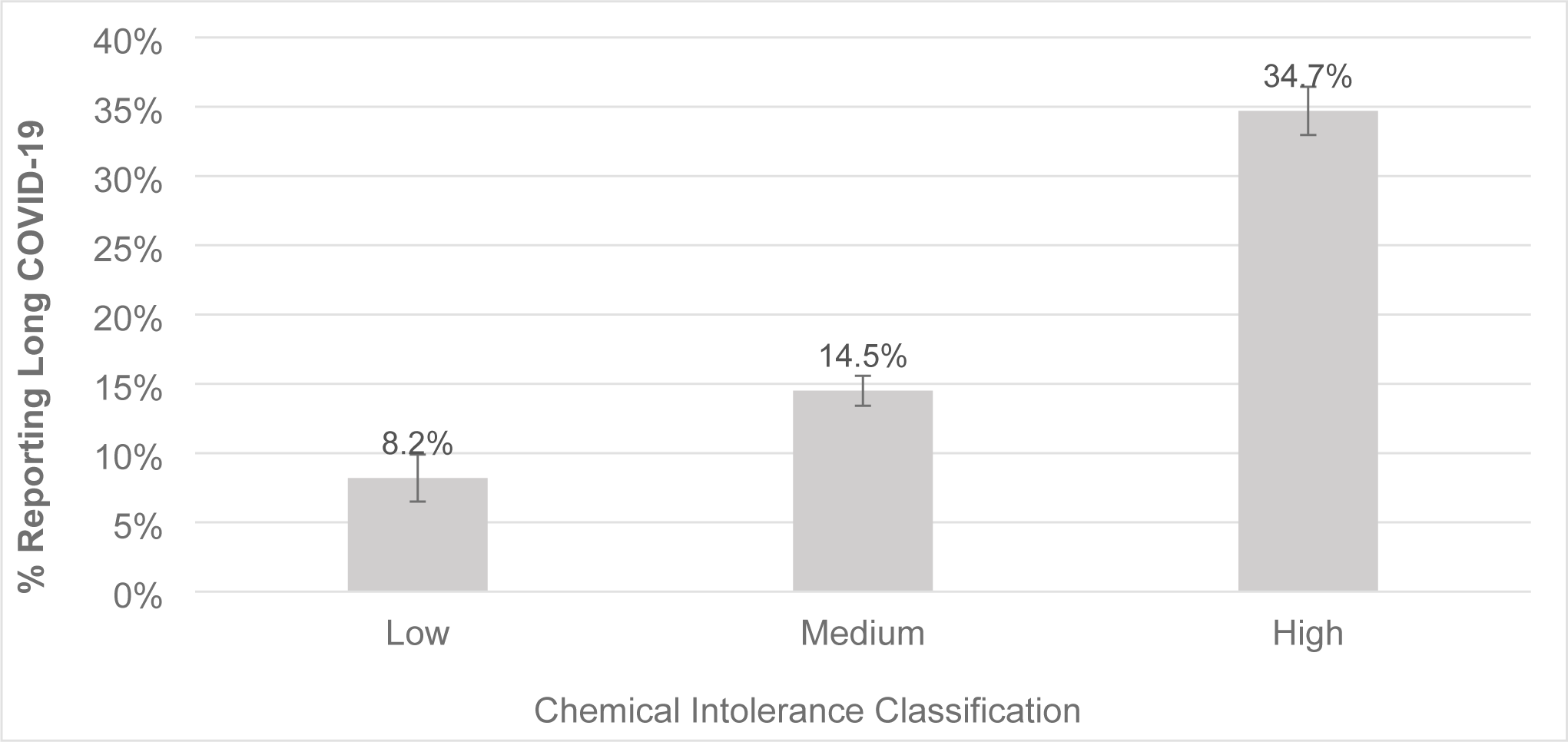
Percent reporting long-COVID-19 by chemical intolerance classification.

A significantly higher percentage of Long COVID-19 is reported by those with the highest level of CI compared to the mid-level (p<.01) or low-level CI (p<.001).

Hispanic or Latino respondents had 1.6 times the odds of reporting Long COVID-19 than non-Hispanic or Latino respondents (95% confidence interval 1.2 – 2.1). Using White/Caucasian as a base, Black/African American respondents had 1.7 times the odds of reporting Long COVID-19 than White/Caucasian respondents (95% confidence interval 1.2 – 2.5). The logistic model demonstrated good model fit (lack of fit P x^2^ = 0.07).

Table 4 summarizes the three aforementioned logistic models. In Summary, those in the High Chemical Intolerance class reported a greater COVID-19 prevalence, severity, and long COVID-19 than those in the Medium and Low CI classes. These associations were independent of race, ethnicity, income, age, and gender. There were significantly increased odds of severity for females and those over 45 years old. The only significant racial coefficient was the comparison between Asian and Whites in the 8+ severity cutoff model, where Asian respondents were least likely to have severe symptoms compared to White respondents (OR = 0.60)

**Table 4.** Summary of the Odds ratios for the three logistic regression models.

|  | Model Odds Ratios |  |  |
| --- | --- | --- | --- |
|  | Prevalence | Severity 8+ | Long COVID-19 |
| High CI vs. Low CI | 3.3*** | 5.0*** | 7.7*** |
| Mid CI vs. Low CI | 1.8*** | 2.2*** | 2.7*** |
| Hispanic / Latino | 1.6*** | 0.7*** | 1.6*** |
| Older than 60 | 0.4*** | ns | ns |
| Female | ns | 1.3*** | ns |
| Race (Black) | 0.7*** | ns | 1.7*** |
| Race (Asian) | ns | 0.5 | ns |
| Race (Black) x High CI interaction<sup>a</sup> | 2.2*** | ns | ns |

<sup>a</sup> Black/African American people with High CI
\*\*\*P < .0001; ns = not significant

Black/African American individuals had an overall lower prevalence of COVID-19 than NHW in this survey, but African American respondents with high CI reported 2.2 greater odds of reporting COVID-19 prevalence. Further, Black, African American respondents had a greater odds of increased symptom severity.

### 3.5 Reactions to the COVID-19 Vaccine, Hypothesis 5

From table 2, compared to the low CI group, those in the High CI group were less likely to have received a COVID-19 vaccination, and to report a significantly greater percentage of moderate and/or severe symptoms (p < .001) (See Figure 6).

**Figure 6.**
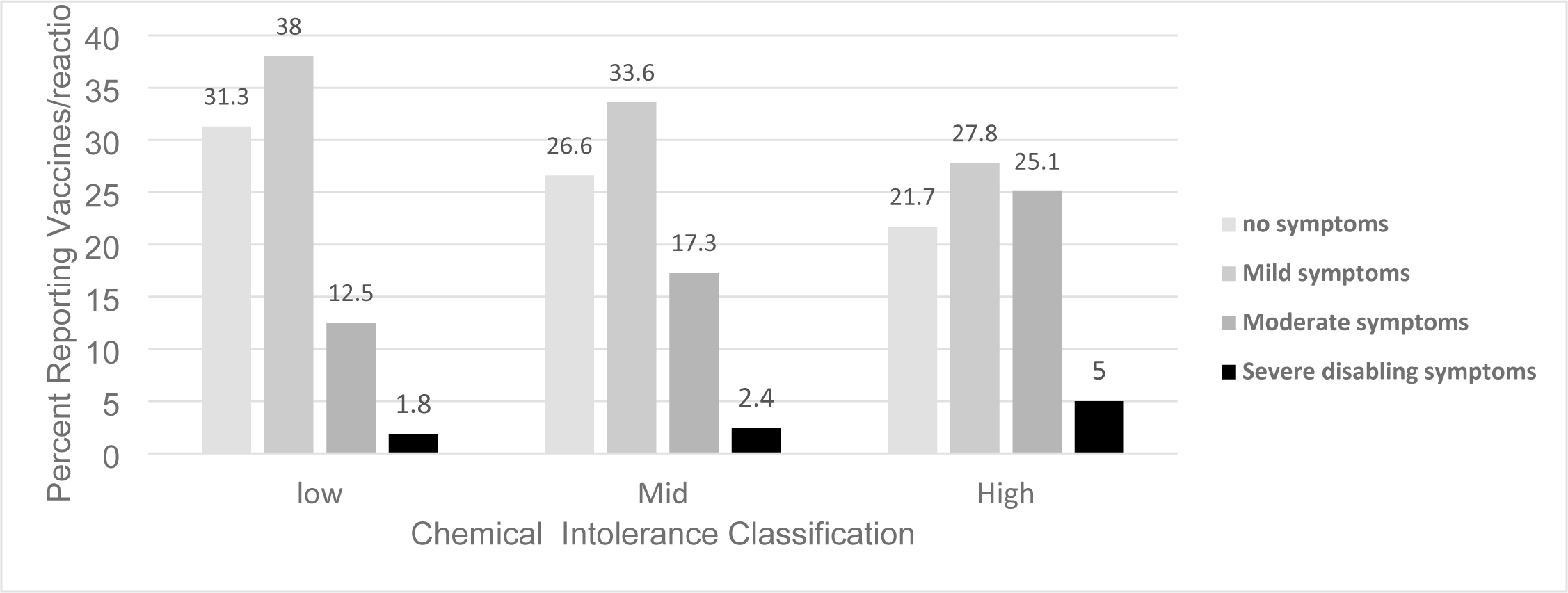
Percent reporting vaccines and vaccine reactions by chemical intolerance group.

## Discussion

This study found that those with CI reported significantly higher rates of COVID-19 compared to those without CI. They also report significantly greater COVID-19 symptom severity and higher rates of long COVID-19. One plausible explanation for these findings is that those with CI have a greater number of preexisting medical conditions than those with low CI [33]. This greater disease burden may further compromise an already taxed immunity response against viruses [9] and/or invoke differential mast cell activation among those with CI [34,35].

It may also be plausible that being infected with COVID-19 somehow initiates CI. However, the current study has insufficient data to make that determination. There is no evidence in the literature that COVID-19 would initiate CI, this would be a matter for further study.

We found that females report worse COVID-19 symptom severity than males. However, this is contrary to other reports indicating that males have worse COVID-19 sympomatology than females [36], however, CI had not been assessed in those studies, so its plausible that CI was a confounder of the prior reports of sex differences in COVID-19 syptomatology. On the otherhand, females have consistently been reporteed to have higher CI than males (11, 22, 37). This suggests a potential subgroup connection between CI and COVID-19-19 severity in females, but our analysis did not find any signficant gender interctions with the COVID-19-19 outcome variables.

We found ethnic and racial group differences related to COVID-19 prevalance, COVID-19 reaction severity, and long COVID-19 status. Compared to NHWs, Hispanic individuals reported increased COVID-19 infection and long COVID-19 prevalance. Hispanic individuals were 1.6 times more likely to report COVID-19 than NHWs. This is highly consistent with prior research using the Household Pulse Survey showing similar results [38]. Kreutzer et al., (1999) report Hispanic ethnicity was associated with physician-diagnosed multiple chemical sensitivity (MCS)(OR = 1.82, 95% confidence interval (CI) 1.21-2.73) [21]. Concomitantly, we also found that Hispanic individuals reported higher CI rates than NHW (49% vs. 29% respectively) (P<.0001)

Given known racial disparities in occupational and other environmental toxic exposures [39], it is plausible to suspect that exposure to toxins may at least partially explain the disproportionate impact of COVID-19 among racial and ethnic minoritized populations [40].

Wong et al., (2022) [40] used the United States Environmental Protection Agency’s Risk-Screening Environmental Indicators (RSEI) Model US EPA(2021) to determine if environmental toxins mediate the association between COVID-19 hospitalizations and race and ethnicity. The RSEI is a measure that considers the amount of toxic chemicals released, the route of exposure, individual chemical toxicity, and the number of people potentially exposed. Higher scores indicate higher risk of toxic exposure [41]. They report that higher RSEI scores were associated with increased COVID-19 hospitalization rates and that all racial and ethnic minority groups had higher odds of hospitalization compared with the non-Hispanic White group. They showed that RSEI decile scores mediated the racial and ethnic COVID-19 hospitalization disparities among Hispanic and non-Hispanic Black individuals [40].

Disparities among Hispanic Women might also be accounted for by marked differences in occupational exposures to pesticides and cleaning agents [42]. According to the Economic Policy Instutite, women make up the vast majority of domestic house cleaning workers, and 63% of those womwn are Hispanic [40,43). Domestic housecleaners are exposed to a myriad of chemical cleaners. Future research would further investigate the connection between occupational exposures, CI, COVID-19-risk, and ethnicity.

In this study, we found that Black/African American respondents reported an increased risk for Long COVID-19 despite having a lower COVID-19 prevalance. However, taking into acount the signficant interaction term, we found that Blacks with High CI are at twice the risk of reporting having COVID-19. This is consistent with the literature showing that African Americans and Hispanics are about twice as likely to need to stay in the hospital due to COVID-19 than non-Hispanic whites [44].

Individuals of Asian race reported a lower COVID-19 severity score than the other ethnic groups. This may be due to an increased willingness among the Asian culture to take greater precaution against infectious disease (e.g., mask wearing) than other cultures [45]. The relatively low number of Asian respondents in this study precluded investigating CI subset interactions between the COVID-19 outcomes.

Another plausible cultural explanation of lower COVID-19 severity scores among Asian individuals may be due to the fact that Asian individuals tend to demonstrate lower extreme response scores than other cultures—believing it is more important to be modest and respond cautiously (46-48). Studies of cross-cultural differences in response styles show that ordinal response formats (e.g., Likert scales) differ across cultures. Asian cultures tend to choose more middle-level items on Likert scales than other cultures [49–50].

### Biological plausibility

In prior work we have demonstrated a strong association between Chemical Intolerance and mast cell proliferation. Approximately 50% of patients with mast cell activation syndrome (MCAS) also have CI, with MCAS patients having similar symptoms as those with CI [34,35]. This suggests that those with CI may more readily respond to environmental insults with greater and a more sustained mast cell immune response. This may include responses to viruses themselves or to vaccinations. Indeed, we demonstrated that those with High CI in this study reported significantly more severe adverse reactions to the COVID-19 vaccine.

### Indoor air and COVID-19

Several studies indicate that proper indoor air ventilation can mitigate the risk of COVID-19 [51,52] However, the COVID-19 quarantine may have posed more risk for those who are chemically intolerant. The quarantine may have affected indoor air quality (IAQ) by increasing adverse exposures for longer periods of time– enduring higher levels of indoor air pollutants than before the pandemic. In the face of poorly ventilated spaces, the risk of infectious disease can be compounded through accumulated volatile organic compound (VOC) by everyday product use including scented cleaning or personal care products, pesticide use, combustion through use of gas stove or burning candles, and plug in air fresheners [53]. It is the collective activities of the individual’s comprising households that affect indoor air quality [54].

To sanitize the indoor environment against COVID-19, the use of these toxic cleaning “air-freshener” products may have increased during the lock down period, therefore potentially triggering CI symptoms. Fragranced cleaning and/or personal care products have been found to be the principal triggers people with CI report [25,55]. This is consistent with other reports in the literature involving the association between poor indoor air quality, systemic, health, and risk of COVID-19 [54,56,57]. This has become a new challenge for physicians and health care providers [58].

### Interventions

In prior work, we demonstrated that substantial improvements in the symptoms of those with CI were achieved through “Environmental House Calls” (EHC) [59,60]. In these EHCs, indoor air quality was assessed in the homes of 37 persons with CI. VOC sources were identified using home air samples analyzed by gas chromatography/ Mass spectrometry (GC/MS). Through a series of home visits, the intervention team taught each person and their families how to reduce their exposures. Pre-post symptom assessments were made. Fragrances were present in every home and the symptoms of those who were able to eliminate household VOC sources including fragrances improved, corresponding with reduced indoor air levels which were measured a second time, post-intervention. Practical advice for physicians and patients on ways to improve indoor air quality and how it affects health are shared in Supplemental materials file.

### Study Limitations

As indicated in the Section 2, the overall study design is observational, and no causality can be established without further research. The survey was conducted via a paid, computerized survey platform (SurveyMonkey). As such, all respondent answers were self-reported and therefore prone to several biases, including social acceptability, honesty, differing interpretations of questions, and recall bias. Payments to participants were small (less than USD 10) and did not constitute “undue influence”. To address both payment and self-report concerns, extensive data quality procedures were employed to remove surveys completed too quickly or illogically.

Although the survey was balanced to reflect state population sizes, participants’ sex, age, race, and education, selection bias in computer-based surveys can be marked. Our computerized surveys suggest under- sampling of Blacks/African American and Hispanics/Latino respondents, both by nearly 50%. Lack of access to the internet, a computer, or a smartphone, as well as language limitations, may have also reduced the generalizability of our findings for low-income and minoriizedt populations [61].

### Conclusion

Prior studies show that individuals at higher risk for COVID-19 infection include the elderly, male sex, those with pre-existing comorbidities (e.g., challenged immunities) and racial/ethnic disparities [62]. The results of this study suggest that those with CI be included in a high risk group. Future investigations could identify various risk subsets. Understanding risk subgroups would be helpful in mounting effective targeted prevention efforts.

## APPENDIX 1

1. Did the overall prevalence rate of reported CI increase from PEI1 to PEI2?

*Hypothesis 1:* The prevalence of CI will be higher in PEI2.

Comparison of PEI1 v PEI2 rates

1. Did those with CI have higher rates of Covid than those without CI?

*Hypothesis 2:* The prevalence of CI will be higher among those with CI.

Have you been diagnosed with, *or do you think you have had* Covid?

Yes, I believe I have had Covid (no test)

Yes, I tested positive for Covid with a home test

Yes, I tested positive for Covid by a health professional

No

I am not sure

1. Are those with CI more severely affected by Covid?

*Hypothesis 3:* Those with CI will be more severely affected by Covid.

If yes, please rate the severity of your Covid symptoms on a 0-10 scale.

[0 = no symptoms] [5 = moderate symptoms] [10 = disabling symptoms]

0 1 2 3 4 5 6 7 8 9 10

1. Are those with CI more likely to experience Long Covid?

*Hypothesis 4:* The prevalence of CI will be higher among those with CI.

Did your Covid symptoms last for more than 12 weeks (long Covid)?

Yes

No

1. Are those with CI more likely to experience reactions to the Covid vaccine?

*Hypothesis 5:* The prevalence of CI will be higher among those with CI.

Please rate the severity of your symptoms after a Covid *vaccination(s)*.

N/A – I have not had any Covid vaccinations

None – I have had Covid vaccination(s) and had no reaction to them

Mild symptoms

Moderate symptoms

Severe (disabling symptoms)

